# Cardiovascular disease biomarkers derived from circulating cell-free DNA methylation

**DOI:** 10.1101/2021.11.05.21265870

**Authors:** Rafael R. C. Cuadrat, Adelheid Kratzer, Hector Giral Arnal, Katarzyna Wreczycka, Alexander Blume, Veronika Ebenal, Tiina Mauno, Brendan Osberg, Minoo Moobed, Johannes Hartung, Claudio Seppelt, Denitsa Meteva, Arash Haghikia, David Leistner, Ulf Landmesser, Altuna Akalin

## Abstract

Acute coronary syndromes (ACS) remain a major cause of worldwide mortality. ACS diagnosis is done by a combination of factors, such as electrocardiogram and plasma biomarkers. These biomarkers, however, lack the power to accurately stratify patients into different risk groups. Instead, we used changes in the circulating cell-free DNA (ccfDNA) methylation profiles to estimate the extent of heart injury and the severity of ACS. Our approach relies on the fact that dying cells in acutely damaged tissue release DNA into the blood, causing an increase in the ccfDNA. In addition, each cell type has a distinct DNA methylation profile. We leverage cell type/state specificity of DNA methylation to deconvolute the cell types of origin for ccfDNA and also find DNA methylation-based biomarkers that stratify patient cohorts. The cohorts consisted of healthy subjects, and patients from three ACS conditions: ST-segment elevation myocardial infarction (STEMI), non-ST-segment elevation myocardial infarction (NSTEMI) and unstable angina (UA). We have used two cohorts of patients - discovery, and validation, both consisting of the same conditions. We have sequenced the ccfDNA from the discovery cohort using Whole Bisulfite Genome Sequencing (WBGS), to obtain an unbiased overview of plasma DNA methylation profiles. We have found a total of 1,614 differential methylated regions (DMRs) in the three ACS groups. Many of the regions are associated with genes involved in cardiovascular conditions and inflammation. Using linear models, we were able to narrow down to 254 DMRs significantly associated with ACS severity. The reduced list of DMRs enabled a more accurate stratification of ACS patients. The predictive power of the DMRs was validated in the confirmation cohort using targeted methylation sequencing of the validation cohort.

## Introduction

Despite a marked mortality reduction driven by improved diagnosis and medical care, ischemic heart disease (a.k.a. acute coronary syndromes) remains a leading health burden in the 21st century ^1^. There is an utmost demand for novel diagnostic tools that refine stratification and monitoring of patient care to improve therapy choice. Chest discomfort is the predominant initial symptom of acute coronary syndromes (ACS), but a combination of criteria is required to achieve the diagnosis of acute myocardial infarction. First, patients with persistent electrocardiogram (ECG) abnormalities are defined as ST-segment elevation myocardial infarction (STEMI) patients ^2^. STEMI is a life-threatening episode accounting for around 30% of ACS cases that can lead to ventricular fibrillation and sudden cardiac arrest and requires immediate intervention ^3^. Plasma biomarkers such as cardiac troponin I (TnI) and T (TnT), or creatine kinase isoenzyme MB (CK-MB) determine the extension of myocardial injury and are additionally used for diagnosis of non-ST-segment elevation myocardial infarction (NSTEMI) ^4–7^. Recent advances in cardiac-specific-troponin (cTn) assays such as high sensitive cardiac Troponin (hs-cTn) have increased their sensitivity and diagnostic value ^8^, but this biomarker is cardiomyocyte-specific and allows estimation of cell stress na injury of only cardiac, whereas the effects of severe hypoxia on other cell types present in the ischemic area remain unknown. Last, patients with normal ECG and no rise of myocardial injury markers (hs-cTn) and chest pain at rest are defined as unstable angina (UA). UA patients which have a lower risk of death and may benefit from less invasive strategies within 72h ^9^ of symptom onset The causes of UA can be numerous and independent of a thrombotic event, but patients often get the same invasive intervention as NSTEMI. It is currently unknown how much the diagnosis of UA predicts future myocardial infarction. It would be extremely beneficial to Identify new noninvasive biomarkers that could improve the stratification between thrombotic or non-thrombotic events leading to different types of myocardial Infarction ^10^.

Nucleosome-sized DNA fragments released from dying apoptotic or necrotic cells are defined as ccfDNA, which circulate for a short time in body fluids before they are cleared,mainly, by the liver ^11,12^. Increased concentrations of ccfDNA have been detected in many conditions: several types of cancer ^13–15^, acute and chronic systemic inflammations ^16^, sepsis ^17^, stroke ^18^, and myocardial infarction ^19^, arising as potential biomarkers for many pathologies.

Recently, several studies demonstrated a strong correlation between circulating cell-free DNA and cardiovascular disease risk factors and status ^19–21^. A cardiac-specific methylated region adjacent to the gene FAM101A has been investigated as a possible biomarker for human cardiomyocyte death ^19,22^. One of those studies found significant differences in the methylation of the 6 CpGs in this region on ccfDNA from STEMI patients and sepsis ^19^.

Analysing ccfDNA fragments by next-generation sequencing (NGS) allows detecting different types of cancer signatures (by detecting mutations related to cancer types) ^15^, monitoring transplant rejection (by detecting increased levels of ccfDNA from organ donor) ^23^ and fetal genetic diseases screening ^24^. Besides variant detection, DNA methylation patterns of ccfDNA, enable the quantification of cell death in a specific tissue, leading to a better understanding of the pathophysiological processes ^25,26^.

In this work, we used BS-sequencing to (i) investigate the changes in the proportion of cells originating the ccfDNA on ACS patients (STEMI, NSTEMI and UA), (ii) investigate differentially methylated regions (DMR) from ACS patients compared with healthy controls, as potential biomarkers for ACS patient stratification, and (iii) test a targeted sequencing approach to validate the DMRs in a new cohort of patients with a more cost-effective method.

## Results

### Clinical parameters and biomarkers

For diagnosis and patient characterization, we measure classical biomarkers levels on study participants (Table S1) and the patients were assigned in the ACS groups by a clinician. High-sensitive cardiac TroponinT (hs-cTnT) were mostly undetected in healthy controls, with low levels in UA patients, and higher values in STEMI and NSTEMI (Figure S1 A). As expected, left ventricular ejection fraction (LVEF) is higher in healthy controls (ranging from 64% to 74%) and lower in ACS patients, with some patients from the UA group showing very low values (minimal value of 30%, figure S1 B). For creatine kinases (CK) we see an increase of the levels with the increased severity of ACS (STEMI the higher average, followed by NSTEMI and UA) (figure S1 C-F).

### ccfDNA concentration, bisulfite conversion and whole genome bisulfite sequencing

The total amount of ccfDNA extracted per mL of plasma from the 29 samples ranged from 2.68 ng (healthy control) to 60.65 ng (STEMI) (figure 1A, table S1). All ACS groups showed higher levels of ccfDNA when compared with the healthy control group (Wilcoxon rank-sum test p-values 0.0001 for STEMI, 0.0003 for NSTEMI and 0.0006 for UA), confirming previous findings were cfDNA levels are raised on diseased states. However, total ccfDNA is not statistically different between ACS samples, preventing the differentiation of ACS types solely by ccfDNA levels.

**Figure 1:**
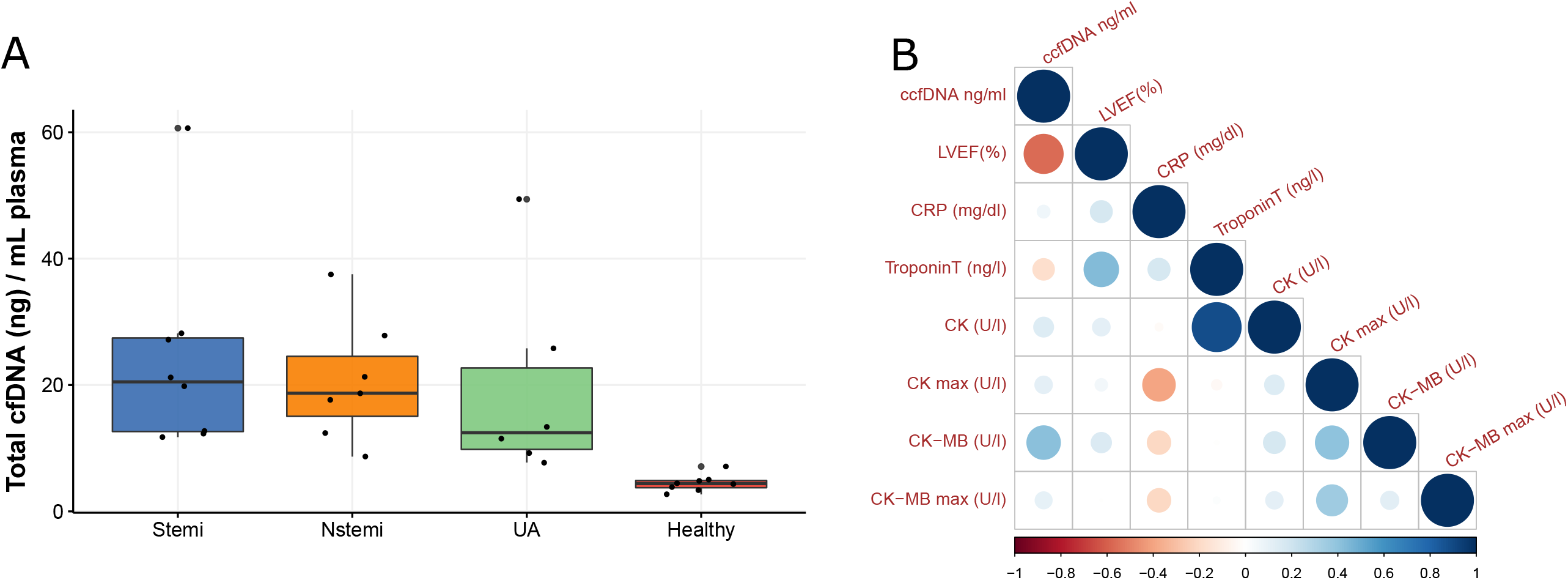
A: Total ccfDNA (ng) per mL of plasma. B: Correlation plot (Pearson correlation) for the classic cardiac biomarkers and ccfDNA concentration (ng per ml of plasma). LVEF(%) - Left ventricular ejection fraction. CRP - C reactive protein. Troponin T - Troponin T high-sensitive. CK - Creatinine Kinase. CK max - maximum value of CK measured. CK-MB - Creatinine Kinase isoform MB. CK-MB max - maximum value of CK-MB measured.

Our quality control analysis showed that we have a good bisulfite (BS) conversion rate, ranging from 97.92% to 99.63% and the average CpG read coverage ranged from 4.2 sequence reads to 9.1 (figure S2).

### ccfDNA correlation with classic cardiac biomarkers in ACS patients

We checked the correlation (Pearson) between all the classic biomarkers and the concentration of ccfDNA (for ACS patients only). ccfDNA concentration in plasma was inversely correlated with left ventricular ejection fraction (LVEF) (R^2^ = -0.56), and, unexpectedly, weakly inversely correlated to troponin. On the other hand, ccfDNA was weakly positively correlated to CK and CK-max and positively correlated to CK-MB (R^2^ = 0.42) (Figure 1B).

### Changes in cell proportions associated with ccfDNA in ACS patients

We used methylation-based cell type deconvolution to quantify the cell-type composition that gave rise to patient-derived ccfDNA. Briefly, our method leverages cell-type specific methylation patterns in the form of a signature matrix. The deconvolution algorithm makes use of those patterns to find optimal proportions of cell types that might give rise to the observed bulk DNA methylation pattern. We used a signature matrix of CpG methylation tissue-specific derived using the dataset from a methylation atlas ^26^ and additional heart-related cell types (see Methods for details). Overall, ccfDNA associated with blood cells were the most abundant in all samples, with neutrophils being the most abundant, ranging from 21% (in the healthy control sample) to 61% (in NSTEMI sample). Previously, granulocytes were found to be the most abundant in healthy donors (average 32% of the cell composition) ^26^, similar to our results for neutrophils, the most abundant type of granulocytes, with an average of 28% for the healthy group. In our data, in all ACS groups, neutrophils were elevated when compared with the healthy control group. Those cells are known to infiltrate infarcted areas in the first hours after ischemia, playing a major role in the subsequent acute inflammation ^27^. We also could detect increased proportions of CD4+ T cells (in NSTEMI and UA), kidney (all ACS) and heart left ventricle (in NSTEMI) tissue. On the other hand, proportions of monocytes were decreased in STEMI and proportions of natural killer (NK) cells, erythrocyte progenitors and hepatocytes were reduced in all ACS (Figure 2).

**Figure 2:**
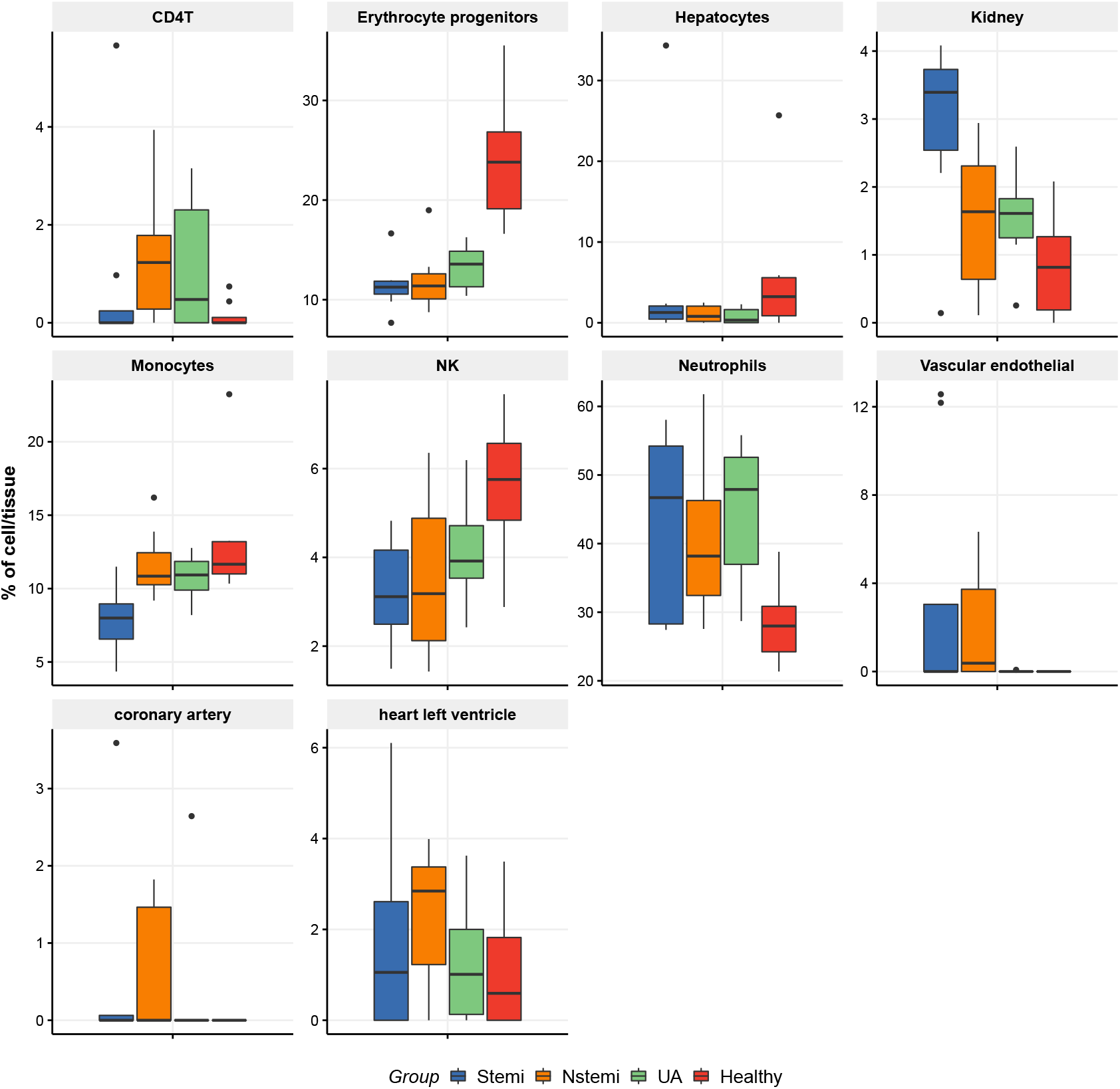
Predicted cell/tissue-type proportions by group (NNLS model) using the extended atlas (only cell/tissue types of interest are shown).

### Identifying and annotating differentially methylated regions (DMRs) in ACS

We conducted a tiled differential methylation analysis using logistic regression based statistical testing as implemented in the Methylkit R package. Using a threshold of minimal 25% difference on methylation and a q-value maximum of 0.01, we identified a total of 688 DMRs in STEMI patients, 388 in NSTEMI patients and 865 in UA. Of those, 486 are STEMI specific, 223 are NSTEMI specific and 684 UA specific. Figure 3A illustrates how the DMRs are shared across the 3 groups. Those disease-specific DMRs are relevant as candidates biomarkers to classify patients in ACS types.

**Figure 3:**
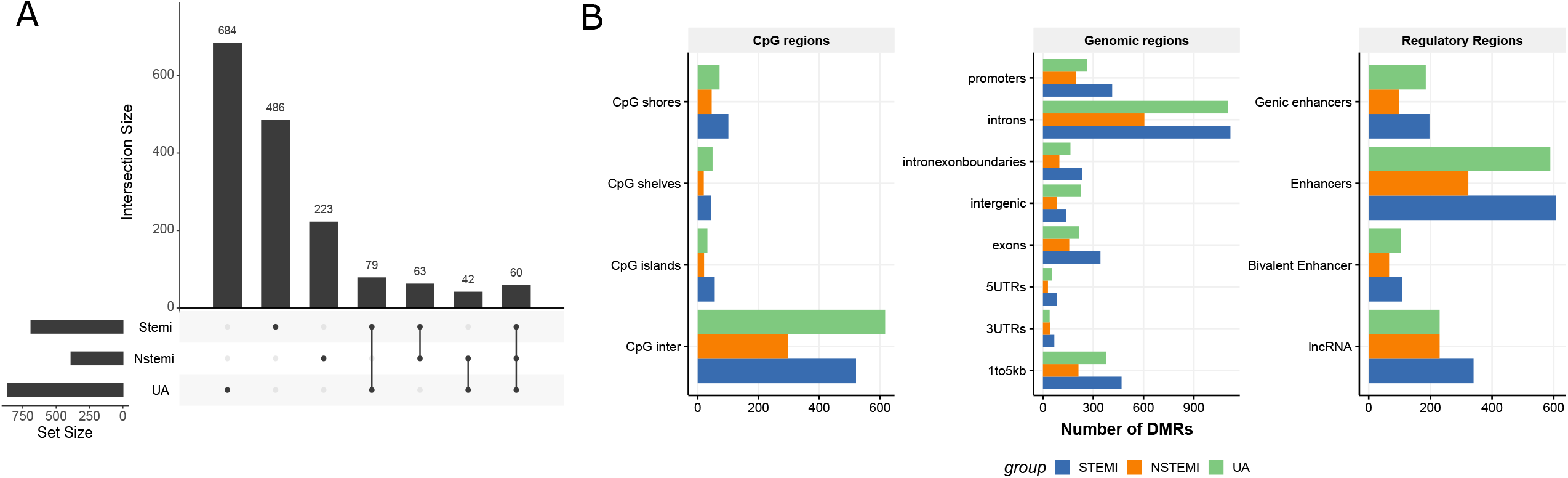
A - DMRs shared across the 3 ACS groups (versus healthy control). B - CpG, genomic and regulatory annotations for DMRs.

Using all identified DMRs, we generated a principal component analysis (PCA). The results show a clear separation of healthy patients from the ACS groups (Figure S3). The UA group is also separated from STEMI and NSTEMI, while these two groups are not clearly separated from each other. However, the ability to differentiate UA and healthy patients from NSTEMI and STEMI is already very useful, as the ECG can be used to further stratify them. We assigned genes that are likely to be regulated by those DMRs using regulatory-region association tool GREAT ^28^. From STEMI DMRs, we found only 4 regions without any gene associated, 118 associated with 1 gene, 565 associated with 2 genes, and 1 associated with more than 3 genes. For NSTEMI, 11 DMRs were not associated with any gene, 64 with one gene, 312 with 2 genes and 1 with more than 3. For UA, 82 genes were not associated with genes, 190 were associated with 1 gene, 592 with 2 genes and 1 with more than 3 genes. Figure S4 shows the number of genes per region for each group, and also the distance from DMRs to the transcription start sites (TSS).

Additionally, we annotated DMRs with CpG island-associated features, genomic features (such as intron/exon,UTRs etc.), Epigenomics Roadmap enhancers and long non-coding RNAs (lncRNA). Most DMRs were in the open sea, out of CpG islands (CpG inter) (564 for STEMI, 329 for NSTEMI and 765 for UA) (Figure 3 B). The genomic annotations show most DMRs on introns (around 60% of all DMRs, figure 3 B) and a large part of it is annotated as enhancers (617 for STEMI, 328 for NSTEMI and 604 for UA) and lncRNAs (91 for STEMI, 43 for NSTEMI and 92 for UA) (Figure 3 B). This points out that differences are mostly driven by cell-type specific regulatory regions.

In addition, for the genes associated with DMRs, we did a gene set enrichment test (using the curated canonical MSigDB C2 pathways, binomial test), finding pathways involved in the regulation of immune response, homeostasis and phagocytosis enriched for the 3 ACS groups (Figure 4A). For STEMI, the most significantly enriched pathway was “Genes involved in hemostasis” followed by “Leukocyte transendothelial migration” (adjusted p-values 0.0003 and 0.0004, respectively). The homeostasis gene set contains genes involved in coagulation, a process known to be triggered during ACS events ^29^. For NSTEMI, we could not detect enriched pathways using the conservative threshold adjusted p-value of 0.05. The lowest adjusted p-value was 0.12 (unadjusted p-value 0.003, Regulation of p38-alpha and p38-beta). For UA, the most enriched pathways were “Genes involved in Regulation of IFNA signalling” and “Fc gamma R-mediated phagocytosis” (both adjusted p-values 0.0001). Fcγ receptors (FcγR) are plasma membrane-associated receptors for IgG and the pentraxins C-reactive protein (CRP), a known risk factor for cardiovascular diseases, and its role on inflammation in cardiovascular disorders have been raised ^30^.

**Figure 4:**
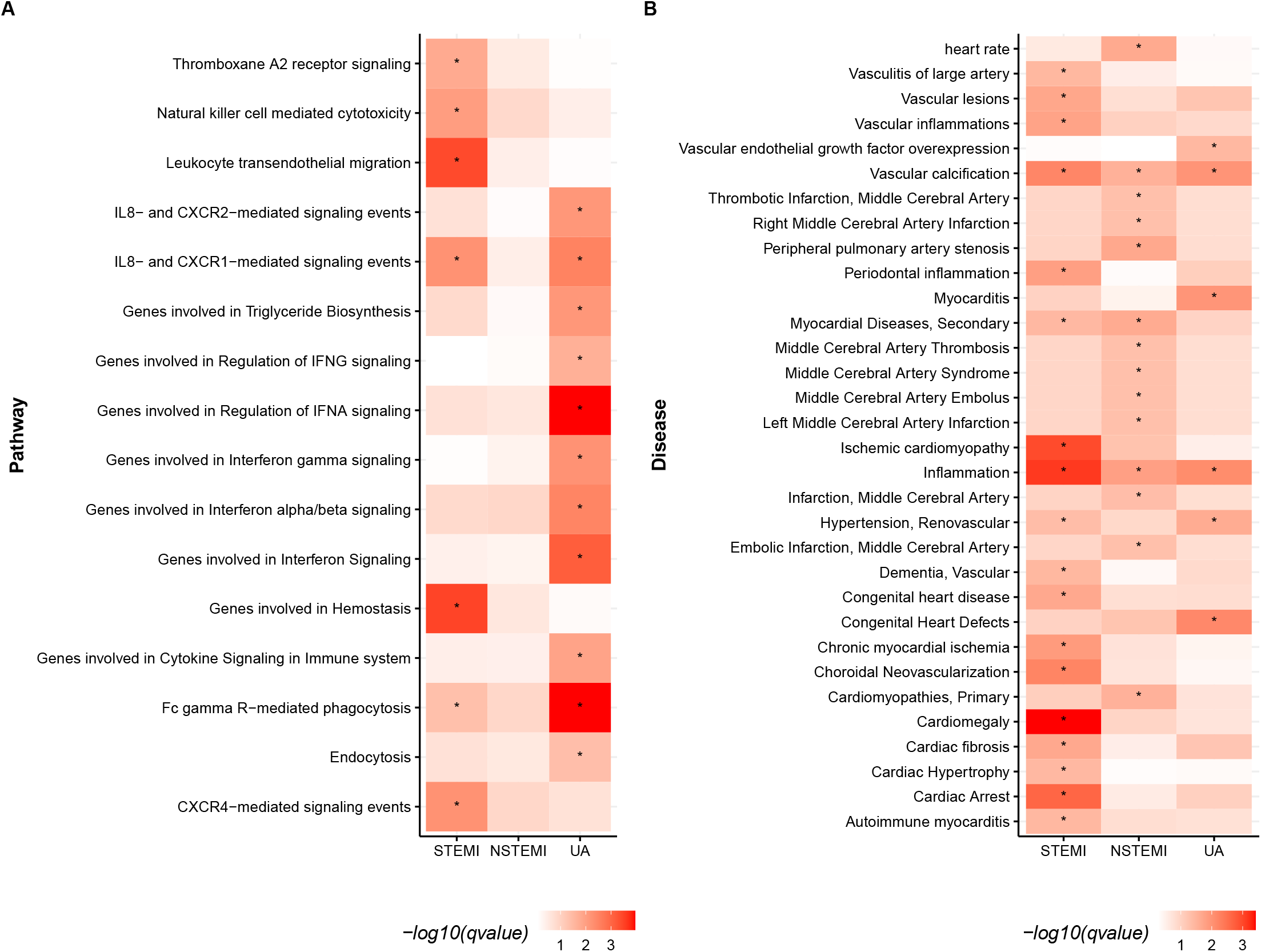
**A - GREAT gene set enrichment for DMRs**. Enriched canonical pathways from MSigDB CP (curated gene sets), with adjusted p-values < 0.05 in at least one group are shown. The colours are representing the adjusted p-values (-log10 based). **B - DisGeNET enrichment analysis showing cardiovascular/inflammatory diseases** w**ith adjusted p-values < 0.05 in at least one group**. The colours are representing the adjusted p-values (-log10 based).

Using DisGeNET ^31^, we assigned the DMR-related genes to diseases. In total, 74.13% of DMRs from STEMI, 72.68% from NSTEMI and 61.62% from UA were annotated with heart-related diseases. The high proportion of DMRs regulating genes involved in heart-related diseases provides us with additional evidence that those DMRs are good biomarkers candidates. Later, we ran a DisGeNET enrichment analysis to check if the DMRs were statistically enriched (q-value < 0.05) for ACS related diseases. We found 505 diseases enriched for STEMI, 253 for NSTEMI and 326 for UA. Filtering by cardiovascular and inflammatory-related diseases, 18 diseases were enriched for STEMI and the most significant was “Cardiomegaly” (q-value 0.00038). For NSTEMI, 14 cardiovascular/inflammatory diseases were enriched, with “Inflammation” the most enriched (q-value 0.01). For UA, only 6 cardiovascular/inflammatory diseases were enriched with “Congenital Heart Defects” the most enriched (q-value 0.007).

Figure 4B shows all the cardiovascular/inflammatory diseases enriched in at least one group, with an adjusted p-value cutoff of 0.05.

### DMRs are associated with disease groups independently of ccfDNA levels in plasma

Because the classic biomarkers cannot be used to easily stratify ACS patients (Figure S1), we decided to further investigate the potential of using DMRs for stratification. While the 1637 discovered DMRs are possible markers for separating each ACS group from healthy individuals (because the differential methylation analysis was done against the control), we decided to try to narrow down a list of DMRs strongly related to the disease group. For that, we ran 2 types of linear models: (i) one model for each classic cardiac biomarker and one for disease severity (as defined on methods) as dependent variables and DMR methylation percentage and ccfDNA concentration as independent variables. (ii) one model for each classic cardiac biomarker (including disease severity) as variable dependent and just ccfDNA concentration as the independent variable.

We found 254 (15.51%) DMRs significantly associated (p-value <= 0.05) with disease groups on the model (i), showing that those DMRs are good candidates for disease stratification. From those, 158 DMRs were annotated as involved in cardiovascular diseases or inflammation using DisGeNET ^31^ and 163 overlap with enhancers from the Roadmap epigenomics project ^32^.

Figure 5A shows the percentage of DMRs significantly associated with biomarkers and disease. Using biomarkers instead of disease groups, we found smaller numbers of DMRs significantly correlated.

**Figure 5:**
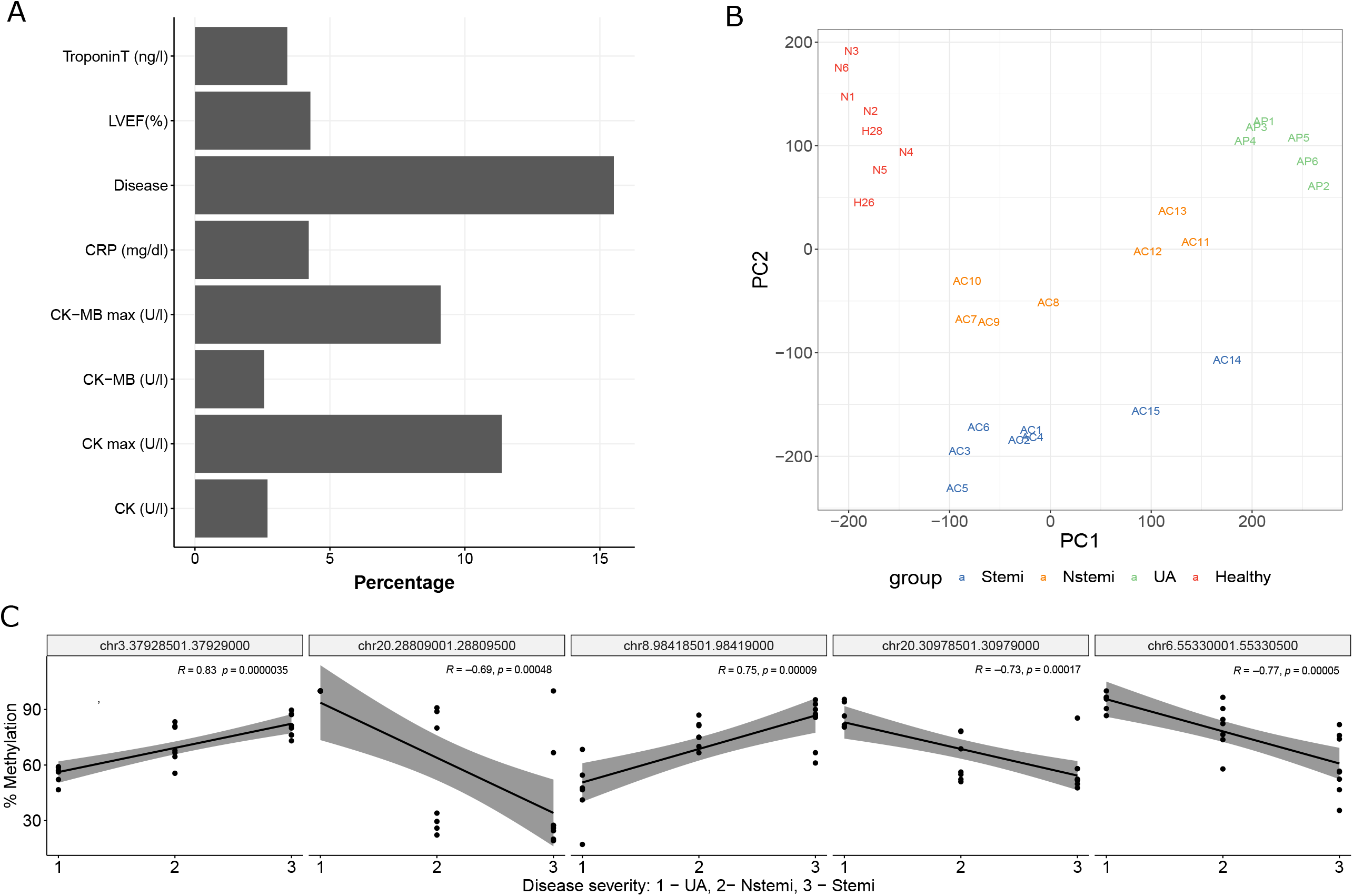
A - Percentage of the discovery DMRs showing significant association with biomarkers and disease group on linear model adjusted by ccfDNA concentration on plasma. B - PCA on methylation levels of 254 DMRs found significantly associated (p<0.05) with disease groups on linear models. C - Correlation plot with the top 5 DMRs most significantly associated with disease groups.

Model (ii) shows only a significant association of ccfDNA with CK-MB(U/l) (p-value 0.046) and not with the disease group. Those results show that ccfDNA alone is not good for patient stratification while using specific DMRs can be useful.

Additionally, we conducted an ANOVA test comparing the models (i) and (ii) to check if adding DMRs to the model is beneficial for prediction of the disease status and biomarkers when compared to a model that only includes ccfDNA concentration on plasma. As expected, because model (ii) mostly shows no associations, for the majority of the DMRs there is a statistical difference between the models, meaning that DMRs are more informative than ccfDNA concentration alone.

Following this, we performed PCA on the methylation levels of the 254 DMRs significantly associated with the disease group (Figure 5B). We can see that with this narrower list of DMRs we have a better stratification and we were able to separate each group. On figure 5C we can see the top 5 DMRs most significantly associated with diseases and its correlation with disease severity.

### DMR validation with target sequencing

For validation of the findings, we tested a target sequencing approach with a small external cohort of patients. We used 2 healthy subjects, 4 STEMI, 3 NSTEMI and 2 UA patients. The participants’ characteristics can be found in table S1. Overall, we targeted 18831 CpGs and around 75% of the targets were covered with at least 5 reads for each sample.

We ran a DMR analysis with the validation samples restricting it to the discovery DMRs (using a q-value cutoff <=0.01). We found that 570 of 681 DMRs for STEMI, 308 of 388 for NSTEMI and 252 of 865 for UA were also differentially methylated on the validation samples. However, the effect sizes (methylation difference) are in general lower in the validation cohort. Those results are preliminary, and, in the future, we will extend the validation cohort and improve the targeting for the shorter list of DMRs that are able to stratify the ACS groups.

## Discussion

In recent years ccfDNA emerged as an interesting new molecular biomarker for diverse diseases ^12,16–18,24^. However, most of the studies investigated the increase in the total amount of ccfDNA, and the correlation of ccfDNA levels with the severity of the diseases. While useful, such an approach is limited because it cannot discern which tissue is being damaged, nor can it measure the level of injury. ccfDNA fragments contain DNA methylation patterns which enable their unique assignment to the cell type where the fragment originated. ccfDNA methylation profiling has been used to determine the cellular contributors of the damaged tissue, for example, in islet transplantation and sepsis ^26^.

In the present study, we present a proof of principle approach, by using unbiased whole genome bisulfite sequencing (WGBS) to *de novo* investigate differential methylation profiles of ccfDNA in three different ACS and a healthy control group. We analysed both the ccfDNA methylation-based cell composition and the changes of methylation in response to the disease state. Additionally, we used a validation cohort to validate the discovered DMRs, using a target sequencing approach. The target sequencing approach greatly reduces the profiling costs, and speed, and makes the method amenable for the possible clinical usage. As previously shown, we found that all ACS groups had increased levels of ccfDNA, when compared to the healthy control group (Figure 1). Interestingly, both MI groups showed slightly higher amounts of ccfDNA than UA. The difference was however not statistically significant, possibly due to the lack of statistical power. A study reported similar, linearly increasing levels of ccfDNA from UA, NSTEMI and STEMI ^33^. It is important to state that the ccfDNA fragments can originate from different cell types, and not only cardiomyocytes.

In fact, our cell composition analysis showed an increased proportion of neutrophils in ACS, a cell type known to be involved in the immune response to MI ^27^. Neutrophiles are the first cell type recruited to the site of injury and contribute different functions in cardiovascular diseases ^34^. During ACS, neutrophiles release extracellular traps. Neutrophil extracellular traps (NETs) are secreted structures formed by decondensed chromatin, histones and neutrophil granular proteins, which have been proposed to contribute to ccfDNA ^35^. These structures have pro-thrombotic activity, and their increased levels are related to larger infarct size and major cardiovascular events ^34,36^.

On the other hand, we observed a decrease in the proportions of erythrocyte progenitors in the ACS groups. Contrary to the other cell types, ccfDNA from those cells does not reflect cell death but rather the process of erythrocyte maturation, when the progenitors lose their nuclei ^26,37^. The decreased proportion of erythrocyte progenitors might reflect a shift in the hematopoietic process, increasing the production/maturation of immune cells while decreasing erythrocyte maturation. Simultaneously, we also observed an increase of CD4+ T-cells, especially in NSTEMI and UA. Cumulative evidence from animal models showed a double role of CD4+ T-cells in ischemia-reperfusion injury and tissue recovery ^38^.

We did not detect ccfDNA from the heart left and right atriums in any of the ACS groups. These results are consistent with the rarity of the atrial infarction, due to the lower oxygen demand of the atrium, when compared to the ventricle. We detected on average 1% of cells from the heart left ventricle in healthy controls, with small increases in ACS, where NSTEMI was on average the most prominent (however the maximum value was observed in one STEMI patient).

Our differential methylation analysis found a total of 1637 DMRs when comparing each of the ACS groups with the healthy controls (Figure 4). Those DMRs clearly separated the ACS groups from healthy controls; however, they lacked the power to accurately separate the STEMI from NSTEMI patients (figure S3). Additionally, we have employed linear modeling to find DMRs where the methylation levels are significantly changed between the different disease groups. This procedure resulted with a set of 254 DMRs, and significantly improved the disease patient stratification (figure 9). The reduction in the number of DMRs which are necessary for accurate patient stratification reduces the sample preparation costs and implies a possible clinical application. Interestingly, 96 of those DMRs are currently not known to be involved in cardiometabolic diseases or inflammation. Using a targeted sequencing approach, we were able to cover ∼75% of the targeted regions and validate ∼70% of the DMRs. This percentage of success on targeted sequencing is expected for the approach used.

While investigating the DMRs, we observed that the majority of ACS related DMRs are located in intronic regions. The intronic DNA methylation events were also previously associated with the etiology of dilated cardiomyopathy ^39^ and coronary artery disease ^40^.

It is of utmost importance to mention that the methylation pattern of the released ccfDNA goes beyond the cell death of cardiomyocytes. It highlights that multiple cell types contribute to ccfDNA, through different biological processes in a disease like acute coronary syndrome. Through the ccfDNA methylation signatures, we are measuring information about a whole spectrum of cellular functions related to acute heart disease, with the final goal of improving both the patient stratification and long term outcome prediction. Usage of ccfDNA methylation as biomarkers is, however, not limited to acute events, but could be used for measuring disease progression in chronic conditions, such as coronary artery events and heart failure. This utility merits the expansion of ccfDNA methylation biomarkers into larger clinical investigation efforts.

## Conclusions

Here, we show the potential of using a set of ccfDNA DMRs for ACS patients stratification. By using 245 DMRs we were able to separate the groups without any other cardiac biomarker. We were able to apply a targeted sequencing approach using our identified DMRs in order to validate our findings in the second cohort of patients in a more cost-effective way. In the future, we plan to expand the cohort types and sizes and to improve the targeted sequencing method to make it useful as a non-invasive tool in clinical settings.

## Methods

### Patient Recruitment and Sample Collection

A total of 29 individuals were recruited for the first discovery cohort. This group included 8 healthy individuals (control), 8 STEMI patients, 7 NSTEMI patients and 6 UA patients, with an average age of 61.5 ranging from 38 to 84. 28 individuals are men and just one healthy control woman.

For our validation using a target sequencing approach, we used a cohort of 2 healthy subjects, 4 STEMI, 3 NSTEMI and 2 UA patients (Table S1).

All included patients gave their consent after being fully informed about the study and its purpose.

Fresh blood samples from patients and healthy controls were collected into Na-Citrate tubes (BD), centrifuged at 1200g at room temperature (RT) for 10min in a swing-bucket centrifuge. Supernatant (SN) was transferred to a 15mL Falcon tube (Greiner) and the tube spun again at 1200g for 10 min.

Double-centrifuged plasma was then transferred into a screw cap plasma collection tube (3mL Kisker) and immediately frozen at -80C until ccfDNA extraction.

STEMI patients were immediately submitted to PCI after confirmed ST-elevation via electrocardiogram (ECG) and parallel troponin measurement.

NSTEMI are patients that show no ST elevation in the ECG and were tested for hsTroponin T using two time points and showed either a 15-20% troponin rise or a level >52ng/mL and were submitted to the catheter lab for PCI within a maximum of 72h.

Unstable Angina Pectoris patients do not show a rise in troponin, basically have a normal troponin base level, but suffer from chest pain at rest and were also submitted for PCI below 72h of admission. All subjects gave their written consent after being fully informed about the study and its purpose before the blood was drawn.

Patient characteristics, including age, disease status, concentration and the total amount of cfDNA obtained, and cardiac biomarkers are described in Table S1.

### Measurement of Clinical Parameters

All clinical parameters were measured through Labor Berlin using standardized assays and procedures.

### ccfDNA extraction

Circulating cell-free DNA (ccfDNA) was extracted from freshly thawed double centrifuged (1200g at RT) citrate-plasma using the Qiagen QIAamp Circulating Nucleic Acid Kit (Cat. #55114)and collected using ultrapure nuclease-free water in a total volume of 50μL and quantified using a Qubit fluorometer 2.0 and the Qubit dsDNA HS Assay kit (Cat. # Q32854). Fragment size is confirmed and validated using Agilent Tape Station 2200 and the ccfDNA screentape or the hsD1000 screentape.

### Whole-genome bisulfite sequencing (WGBS)

Isolated ccfDNA was sent to Novogene and rechecked for quality and concentration by the company. Bisulfite conversion and sequencing was performed using their low-input BS-seq (PBAT) protocol. Data were analyzed using previously in-house programmed R software packages (ref. Altuna methylkit).

### Targeted Methylation Sequencing

Input amount and pulldown were optimized using sheared genomic DNA (ZymoResearch Cat. # D5014). A total of 10ng of isolated cell-free DNA per sample, quantified and checked for quality, was used as starting material. For enzymatic conversion as an alternative to bisulfite conversion, we applied the NEBNext Enzymatic Methyl-seq Module (Cat. #E7120S) together with the Nonacus Cell3TMTarget: Library Preparation kit. Up to 8 libraries of individually adapter-tagged samples were pooled to a total amount of 1μg for probe hybridization and capture enrichment. Captured library DNA is quantified and quality checked using Qubit Fluorometer and qPCR as well Agilent 2200 TapeStation with High Sensitivity D1000 reagents and screentape. Samples are loaded onto a SP flow cell and sequenced using the Novaseq 6000 platform acquiring 400 million reads.

### Sequencing quality control, alignment to the reference and methylation calling

Raw reads were processed using the PiGs BSseq pipeline ^41^. First, they were checked for quality with FastQC ^42^ (version 0.11.8). Then, the read sequences were trimmed using Trim Galore wrapper ^43^ (version 0.6.4, cutadapt version 2.6, with arguments “--clip_R2 19 -- three_prime_clip_R2 30” “--clip_R1 9” “--three_prime_clip_R1 30”), removing Illumina adaptors and sequences with quality Phred score smaller than 20. For the WGBS samples (discovery), the bwa-meth (version 0.2.2) ^44^ was used to map reads to reference the human genome (hg38), Picard MarkDuplicates ^45^ (version 2.20.4) for deduplication and methylDackel software package ^46^ (version 0.5.1, with arguments --minDepth 1 -q 5 -p 5) for methylation calling. For the targeted sequencing (validation), the reads were aligned using Bismark (version 0.20.1, with arguments -N 0 -L 20 --pbat) and deduplicated using samblaster (version 0.1.24) and methylKit (version 1.16.0) ^47^ was used for methylation calling.

### Cell-type/tissue deconvolution

The deconvR package was created for the deconvolution of cfDNA to origin cell types. The package was made using R 4.0.2 on RStudio. The deconvolute function within deconvR contains four choices of models. The four choices are non-negative least squares (implemented using the nnls package v.1.4), support vector regression (implemented using the e1071 package v.1.7.4), quadratic programming (implemented using the quadprog package v.1.5.8), and robust linear regression (implemented using the MASS package) ^48^.

Using the simulateCellMix from the deconvR package (https://github.com/BIMSBbioinfo/deconvR), we simulated a dataset containing 1000 mixed samples, using a reference atlas containing the methylation signatures of 25 cell types ^26^. This dataset was deconvoluted using 4 different models using the deconvR package, and these predictions were then compared to the data frame containing the actual cell-type proportions, to calculate the accuracies and weaknesses of each model. The lowest root-mean-square error (RMSE) value was obtained with the NNLS model, followed by the RLM model.

Sample data was first mapped by genomic loci to CpG probe IDs. This was done using the BSmeth2Probe function in deconvR. The genetic locations of CpG probe IDs were specified according to the Illumina Infinium MethylationEPIC v1.0 B5 Manifest File ^49^. The returned table mapping CpG probe IDs to methylation values was then used for the deconvolution. The model NNLS was used for the deconvolution as previous analysis on simulated data had found it to be the most accurate. A comprehensive human methylome reference atlas provided the CpG methylation values used for training the model. This atlas is composed of 28 human cell types (as columns) and roughly 13000 CpG loci (as rows), identified by their Illumina Probe ID ^50^. The trained model then applied to the mapped data resulted in an estimation of origin cell-type proportions of the samples.

The comprehensive array-based human cell-type methylation atlas as generated by Moss et al (2018) ^26^ was extended to include additional heart-related tissues using the pre-filtered full reference methylation atlas and pre-processing scripts provided through the associated GitHub repository (https://github.com/nloyfer/meth_atlas). EPIC Methylation-array data for the right atrium auricular region (n=2, ENCSR517JQA and ENCSR280LMY), heart left ventricle (n=2, ENCSR515ZCU and ENCSR190PQG) and the coronary artery (n=2, ENCSR688OHW and ENCSR582BMR) were acquired from the ENCODE portal ^51^. The methylation-array idat files were renamed to respective red/green channels and then pre-processed using the meth_atlas process_array.R script to normalise the data using an arbitrary reference sample, filter by p-value, sex chromosomes and bead number, and finally remove SNPs and non-CpG sites. An extended methylation atlas was constructed by merging the pre-processed ENCODE methylation data with the reference methylation atlas based on CpG probe id, then CpGs with missing values or sites where the row-wise variance was below 0.1% were omitted and replicates were pooled per tissue. The feature selection of tissue-specific CpGs from the extended atlas was done in two ways. First, the 200 most hypermethylated and hypermethylated CpGs were selected as described by Moss and colleagues ^26^ with minor adjustments. The methylation atlas was scaled by dividing each row of the atlas by the summed methylation values of the respective row, then for each cell type, the top 100 hypermethylated CpGs with the highest scaled methylation values were selected and recorded to prevent repeated selection of the same CpGs. This procedure was repeated for the reversed scaled methylation matrix to identify the top 100 hypomethylated CpGs per cell type. Secondly, using the unscaled extended atlas, the dmpFinder function of the minfi R package ^52^ was used to identify for each cell type the 200 most differential CpGs compared to any other tissue. The sets of tissue-specific CpGs were joined and based on those, neighbouring CpGs within a distance of 50 bp and pairwise-specific CpGs were added as explained in Moss et al (2018) ^26^.

### Differential methylation

The differential methylation was called on tiled regions (500 base pairs sliding windows with a step size equal to 500bp) using the R package methylKit (version 1.16.0) ^47^, q-value cutoff 0.01 and minimal methylation difference of 25%.

### DMR annotation

The DMRs were annotated by Genomic Regions Enrichment of Annotations Tool (GREAT)^28^ using the rGREAT package (release 3.12). The model used was the “Basal plus extension”, annotating genes on proximal regions (5 kilobases upstream, 1 kilobase downstream) plus distal (up to 1000 kilobases). We also annotate the DMRs for CpG regions, genomic regions and regulatory regions using the AnnotateR package ^53^ and additionally we annotate enhancer regions using the chromHMM bed files from the Roadmap Epigenomics Project ^32^ (Core 15-state model).

The list of gene symbols obtained was submitted to DisGeNET (v 7.0), a comprehensive platform integrating information on human disease-associated genes and variants ^31^, using its R package disgenet2r (v0.0.9, database = all). The gene symbols obtained from GREAT were converted to Entrez gene ids and used for DisGeNET enrichment analysis (enrichDGN) with R package DOSE (v 3.12) with parameters p-value cutoff < 0.05, adjusted p-value < 0.2, minGSSize = 2, maxGSSize = 500, pAdjustMethdod = BH, background genes = whole human genome). We looked for heart-related disease by searching terms from the disease name and disease class name from DisGeNET, assigning to each DMR the annotation obtained.

### Linear Models of the methylation levels on DMRs

We defined ACS group as a crescent numeric list of severity (UA = 1, NSTEMI = 2, STEMI = 3) and then, using R lm method, we ran 2 types of linear models: (i) one model for each classic cardiac biomarker (including disease severity as defined above) as dependent variables and DMR methylation percentage and ccfDNA concentration as independent variables. (ii) one model for each classic cardiac biomarker (including disease severity) as variable dependent and just ccfDNA concentration as the independent variable.

## Data Availability

All data produced in the present study are available upon reasonable request to the authors

## Acknowledgement

We thank Dr. Vedran Franke for his valuable suggestions. We also would like to acknowledge the assistance of Dr. Tatiana Borodina from the MDC Genomics facility for her support in performing quality control of our samples and our targeted sequencing approach. Additionally, we want to express our gratitude to the patients and healthy subjects for their valuable blood donations supporting our research.

## Authors’ contributions

AA initially conceived and designed the study and planned it with the input from AK, HGA and UL. Patient recruitment was performed by JH, CS, DM, AH and DL. Plasma isolation, ccfDNA extraction, sample quality check and library preparations were done by MM and TM. VE implemented the deconvolution algorithm and applied it on the samples. Initial raw data processing was done by BO, then repeated by KW. KW performed the initial DMR analysis. Cell type specific signatures are extracted by AB. RC combined all the analysis and performed targeted sequencing analysis as well as all downstream analysis. RC, AK, HGA and AA wrote the paper with input from all other authors.

## Availability of data and materials

Data will be submitted to EGA (European Genome Archive)

## Declarations

### Ethics approval and consent to participate

The study has been conducted according to the declaration of Helsinki and was approved by the Berlin State Ethics Committee in Berlin, Germany EA4/122/14 and EA1/270/16).

### Consent for publication

Not applicable.

---

**Figure S1:**
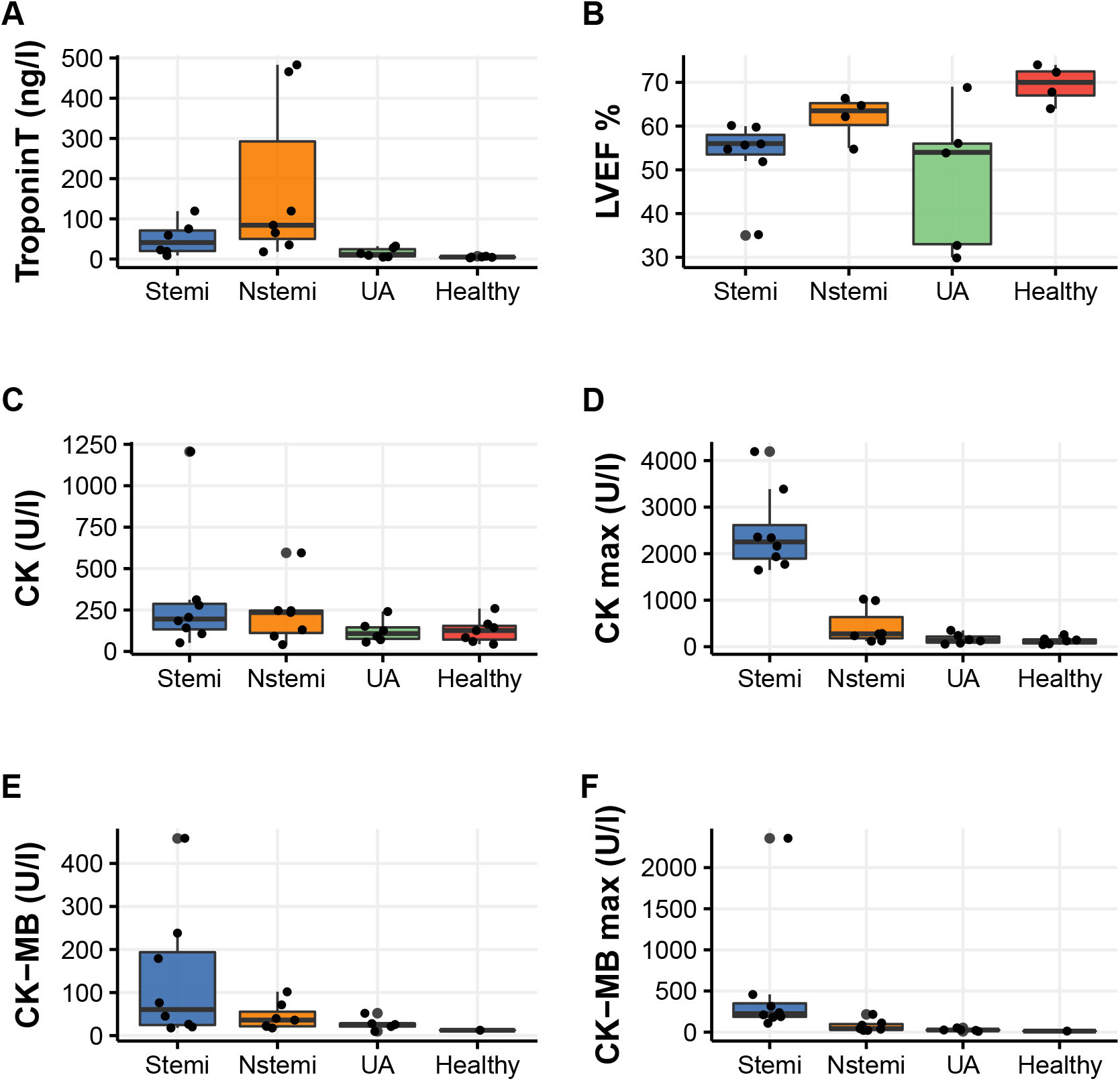
Biomarker values for each experimental group.

**Figure S2:**
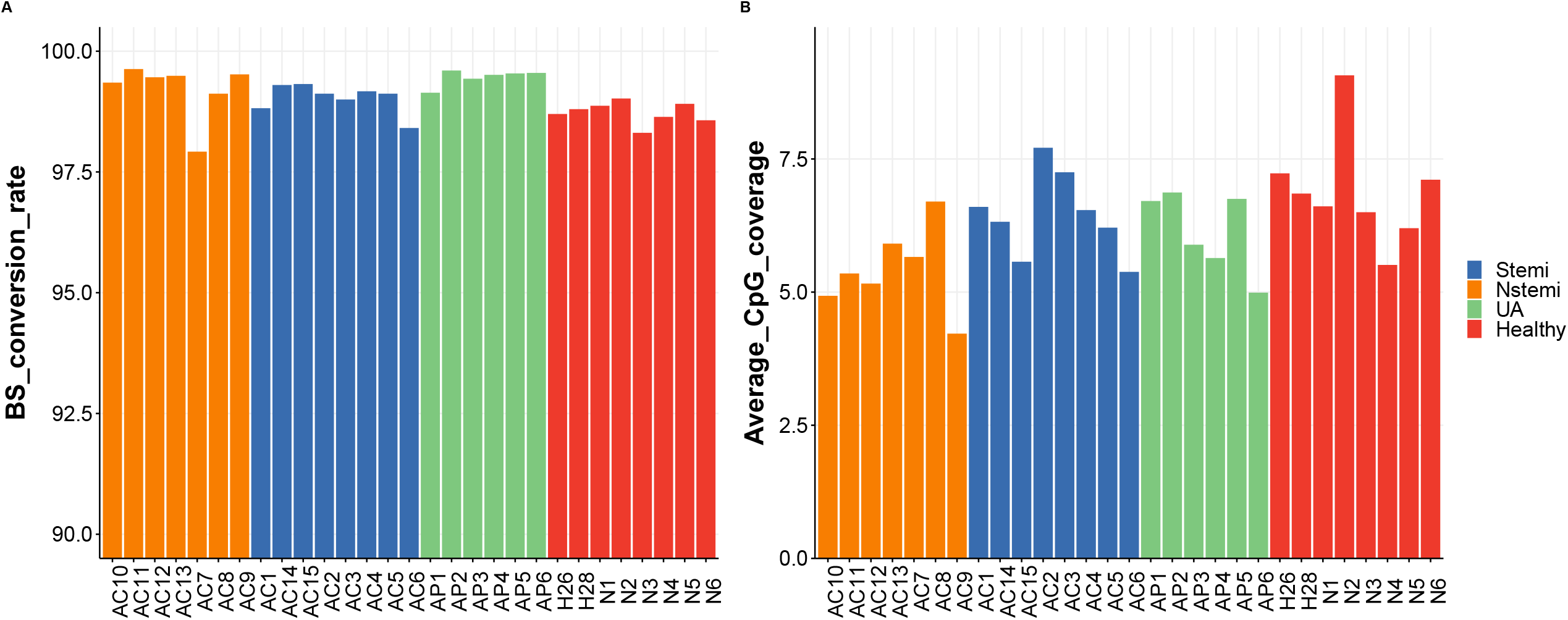
A) BS conversion rate; B) Average CpG coverage for the cfDNA samples.

**Figure S3:**
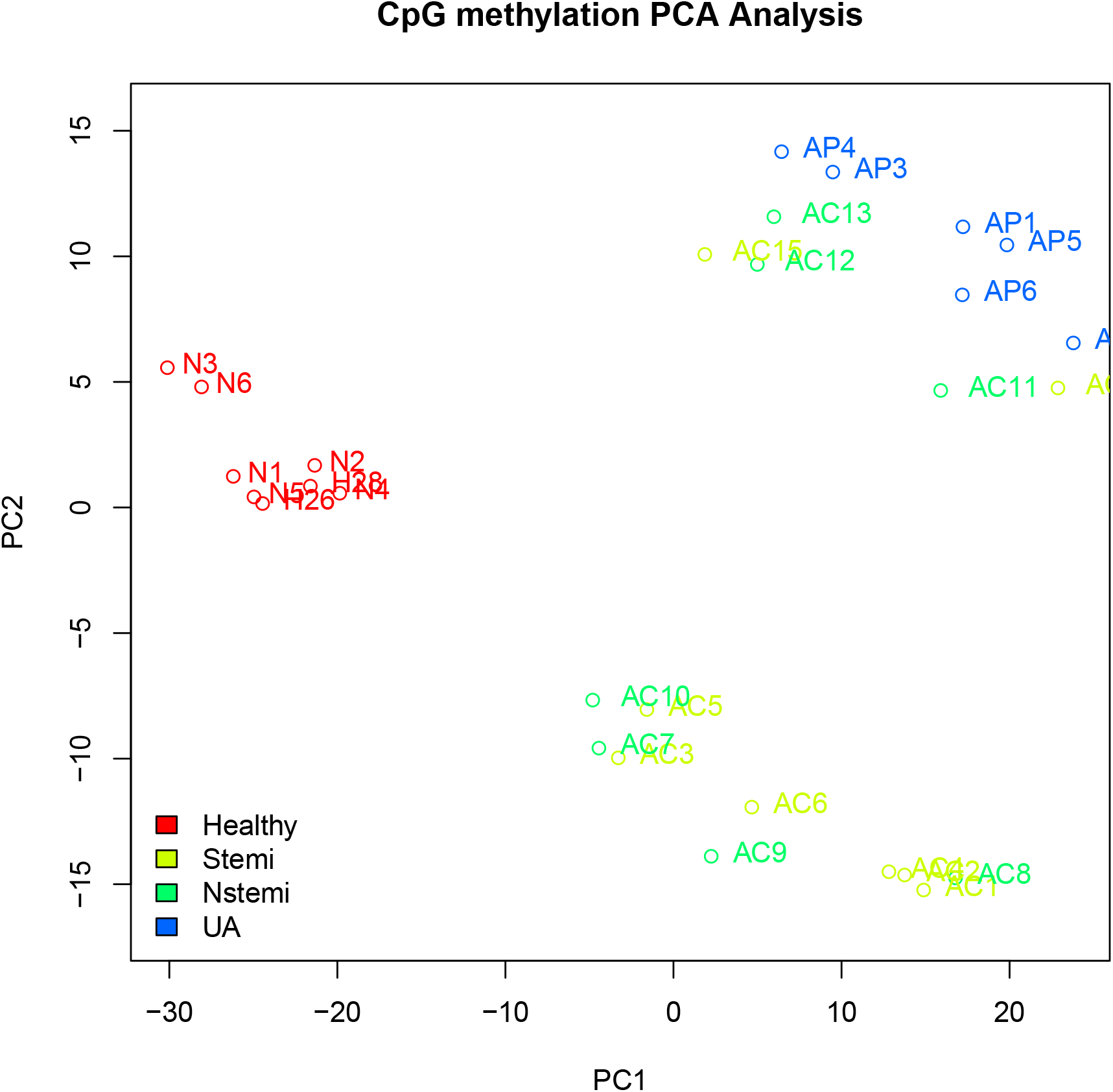
PCA on CpG methylation percentage restricted to DMRs from discovery samples.

**Figure S4:**
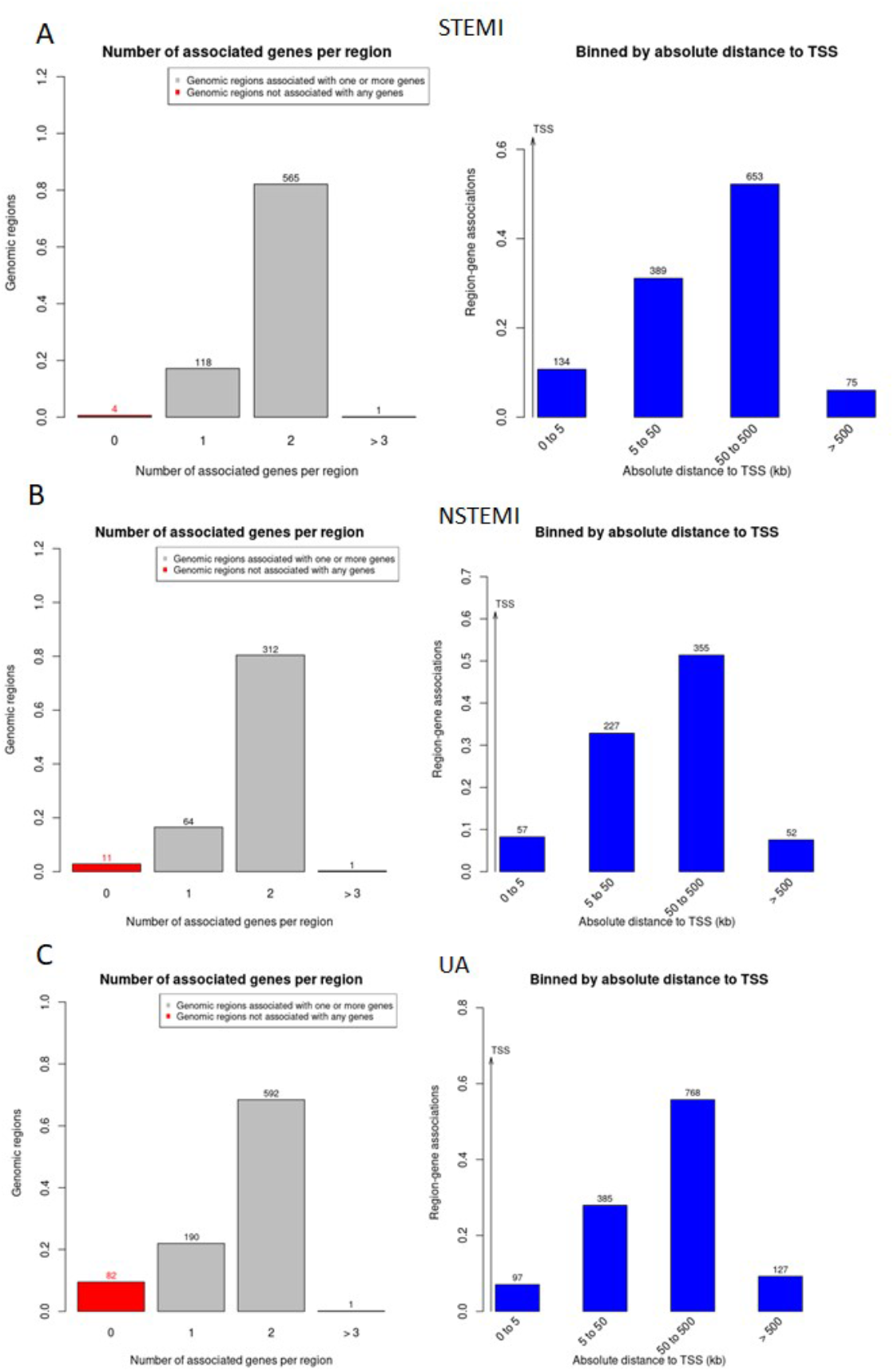
GREAT gene associations and annotations.

